# Founder effect contributes to the unique pattern of SARS-CoV-2 variant B.1.1.519 emergence in Alaska

**DOI:** 10.1101/2022.03.17.22272446

**Authors:** Tracie J. Haan, Lisa K. Smith, Stephanie DeRonde, Elva House, Jacob Zidek, Diana Puhak, Matthew Redlinger, Jayme Parker, Brian M. Barnes, Jason L. Burkhead, Cindy Knall, Eric Bortz, Jack Chen, Devin M. Drown

## Abstract

Alaska is the largest geographic state in the United States with the lowest population density and a mix of urban centers and isolated rural communities. The differences in population dynamics in Alaska from the contiguous United States may have contributed to a unique pattern of emergence and spread of SARS-CoV-2 variants observed in early 2021. Here we examined 2,323 virus genomes from Alaska and 278,635 virus genomes from the contiguous United States collected between the first week of December 2020 through the last week of June 2021. We focused on this timeframe because of the notable emergence and spread of the SARS-CoV-2 lineage B.1.1.519 observed in Alaska. We found that this variant was consistently detected in Alaska from the end of January through June of 2021 with a peak prevalence in April of 77.9% unlike the rest of the United States with a peak prevalence of 4.6%. In Alaska, the earlier emergence of B.1.1.519 coincided with a later peak of Alpha (B.1.1.7) when compared to the rest of the United States. We also observed differences in the composition of lineages and variants over time between the two most populated regions of Alaska. Although there was a modest increase in COVID-19 cases during the peak incidence of B.1.1.519, it is difficult to disentangle how social dynamics conflated changes in COVID-19 during this time. We suggest that the viral characteristics, such as amino acid substitutions in the spike protein, and a founder effect likely contributed to the unique spread of B.1.1.519 in Alaska.

## Introduction

In late 2020 and early 2021, variants of severe acute respiratory syndrome coronavirus 2 (SARS-CoV-2), the causative agent of coronavirus disease 2019 (COVID-19), began emerging worldwide. Variants were found to contain mutations that affected transmissibility, disease severity, diagnostic detection, vaccine efficacy and immune evasiveness compared to the original SARS-CoV-2 virus (Wuhan-1, 2019) lineage (Walensky et al., 2021). Sustained high levels of transmission, variable vaccine and infection-induced immunity, along with geographic and social factors all contribute to the evolution, emergence, and spread of variants in locales across the world.

Alaska is the largest state in the United States by area but has the lowest population density with approximately 1.2 people per square mile on average (US Census, 2020). Population density is highly location dependent across the state with two densely populated and highly interconnected economic regions and many sparsely populated rural areas only accessible by boat or plane. The unique community dynamics in Alaska, paired with its 800 km physical distance from the contiguous United States (hereafter, Lower 48), has the potential to contribute to unique patterns of SARS-CoV-2 variant emergence and spread.

Here, we report on the striking difference in prevalence of SARS-CoV-2 variant B.1.1.519 (Rambaut et al., 2020) between Alaska and the Lower 48 in early 2021. Based on all publicly available genomes outlined in outbreak.info, B.1.1.519 has been detected in at least 72 countries. At its peak, B.1.1.519 comprised around 3% of sequenced cases globally (Mullen et al., 2020) with the highest prevalence observed in Mexico where the lineage was first recognized and comprises 7,959 of 54,886 (14.5%) sequenced genomes (Rodriguez-Maldonado 2021). B.1.1.519 was detected at relatively low levels across the Lower 48 but was never characterized as a variant of concern (VOC), variant of interest (VOI), or variant being monitored (VBM) by the Center for Disease Control and Prevention (CDC). In contrast, the World Health Organization designated B.1.1.519 a variant under monitoring (VUM) on June 20, 2021, implying the potential future risk until November 2021 when Delta became the dominant variant globally (World Health Organization [WHO], 2021). This classification was in part due to the several key mutations B.1.1.519 encodes including amino acid substitutions T478K, P681H, and T732A in the spike (S) protein, that may affect affinity to the SARS-CoV-2 receptor, human angiotensin I converting enzyme-2 (hACE-2). Notably, a study examining the clinical impact of B.1.1.519 infection in Mexico City found patients infected with B.1.1.519 displayed a significant increase in the severity of COVID-19 respiratory tract symptoms relative to patients infected with non-B.1.1.519 variants (Cedro-Tanda et al., 2021).

In this study, we report the emergence, prevalence, and viral traits of SARS-CoV-2 lineages circulating in Alaska from 2,233 sequenced specimens collected between the first week of December 2020 through the last week of June 2021, after which B.1.1.519 was no longer detected. Using SARS-CoV-2 sequence data from the United States available on NCBI GenBank by the CDC and its contracted labs, we explore how the trend in B.1.1.519 and other variants observed in Alaska compared to trends across the Lower 48. These results are used to make predictions about the future of SARS-CoV-2 in Alaska and highlight the importance of continued genomic surveillance for aiding efforts in controlling the pandemic.

## Materials and Methods

### Retrieving SARS-CoV-2 Sequence Data for Alaska

The data presented here were generated as part of the *Alaska SARS-CoV-2 Sequencing Consortium* which is a partnership between the University of Alaska and the Alaska Division of Public Health (AKDPH) with the aim to increase genomic surveillance of variants. On February 14th, 2022, we downloaded 2,323 sequences from samples collected between 2020-11-29 to 2021-06-26 available on GISAID for subsequent analysis (Shu &McCauley, 2017; Elbe &Merret, 2017). Genome sequencing in Alaska is from a non-targeted sample of cases, which is the best available approximation of random samples despite potential disparate coverage across Alaska’s economic regions. We used these data to estimate the prevalence of lineages per week on dates beginning on Sunday.

### Analysis of SARS-CoV-2 Sequence Data for Alaska

Lineages were determined by running sequences through PANGO-v 1.2.124, Pangolin v 3.1.20, and pangoLEARN v 2/2/22, and Scorpio v 0.3.16 (O’Toole et al., 2021). We estimated the prevalence of genomes grouped by the emerging lineages including B.1.617.2 and AY sublineages (Delta), B.1.1.519, B.1.427/429 (Epsilon), B.1.525 (Eta), B.1.617.1 (Kappa), C.37(Lambda), B.1.526 (Iota), B.1.621 (Mu), B.1.351 (Beta), P.1 (Gamma), and B.1.1.7 (Alpha). Genomes that did not fall into these lineages were grouped together into the category ‘Not Emerging Lineage.’ The number of Spike protein amino acid substitutions was determined using Nextclade (v 1.13.2) with ‘bad’ quality genomes, as determined by Nextclade’s algorithm for quality, removed prior to analyses (Aksamentov et al., 2021). Nextclade’s quality control incorporates six individual rules based on missing data, mutations, and amino acid changes as described in the online documentation.

We collected metadata on the number of cases for Alaska from the AKDPH COVID-19 CSV Files Database Cases Data B dataset on 2022-2-14 (AKDPH, 2022). We collected 2020 population estimates for economic regions of Alaska from the Alaska Department of Labor and Workforce Development population estimates page (Alaska Department of Labor and Workforce Development, 2021). Information on the timeline of COVID-19 policies (in terms of restrictions, opening, and vaccines) in Alaska was collected from the Johns Hopkins Coronavirus Resource Center (John Hopkins, 2022)

### SARS-CoV-2 Sequence Data for the Lower 48

On February 14th, 2022, we downloaded metadata available on GenBank for 1,331,799 sequences including Pangolin assignment, Isolate, and GeoLocation for all samples from the United State of America collected between 2020-11-29 to 2021-06-26 sequenced by the CDC. We then filtered out cases from Alaska, Hawaii, and US territories to limit our comparisons to the lower 48 contiguous states. We collected daily case data with seven day rolling averages from the CDC COVID Data Tracker site for the entire United States (CDC, 2022).

### Visualizations and Statistical Analyses

We generated visualizations in RStudio (v 1.4.1106) using packages ggplot2 (v 3.3.5), ggpubr (v 0.4.0), tidyverse (v 1.3.1), and lubridate (v 1.7.10). To test for significant differences in spike glycoprotein amino acid substitutions between emerging lineages or sub-lineages, we used the Wilcoxon test.

## Results and Discussion

### Higher prevalence of B.1.1.519 in Alaska versus the Lower 48

We found a striking difference in the percentage of sequenced cases, a metric that can be used as an estimate of prevalence, assigned to the lineage B.1.1.519 between Alaska (Figure 1B) and the rest of the United States (Figure 1D) in early 2021. While the earliest detection of B.1.1.519 in the United States occurred 27 December 2020 in sequence data from New York, it was not until five weeks later, 4 February 2021, that B.1.1.519 was detected in sequence data from Alaska. By this time in the United States, other VOCs including Alpha (B.1.1.7) and Epsilon (B.1.427/429) had already emerged and were being consistently detected in sequence data. In contrast, Alaska had detected a single case of Alpha and no cases of Epsilon in December of 2020.

**Figure 1.**
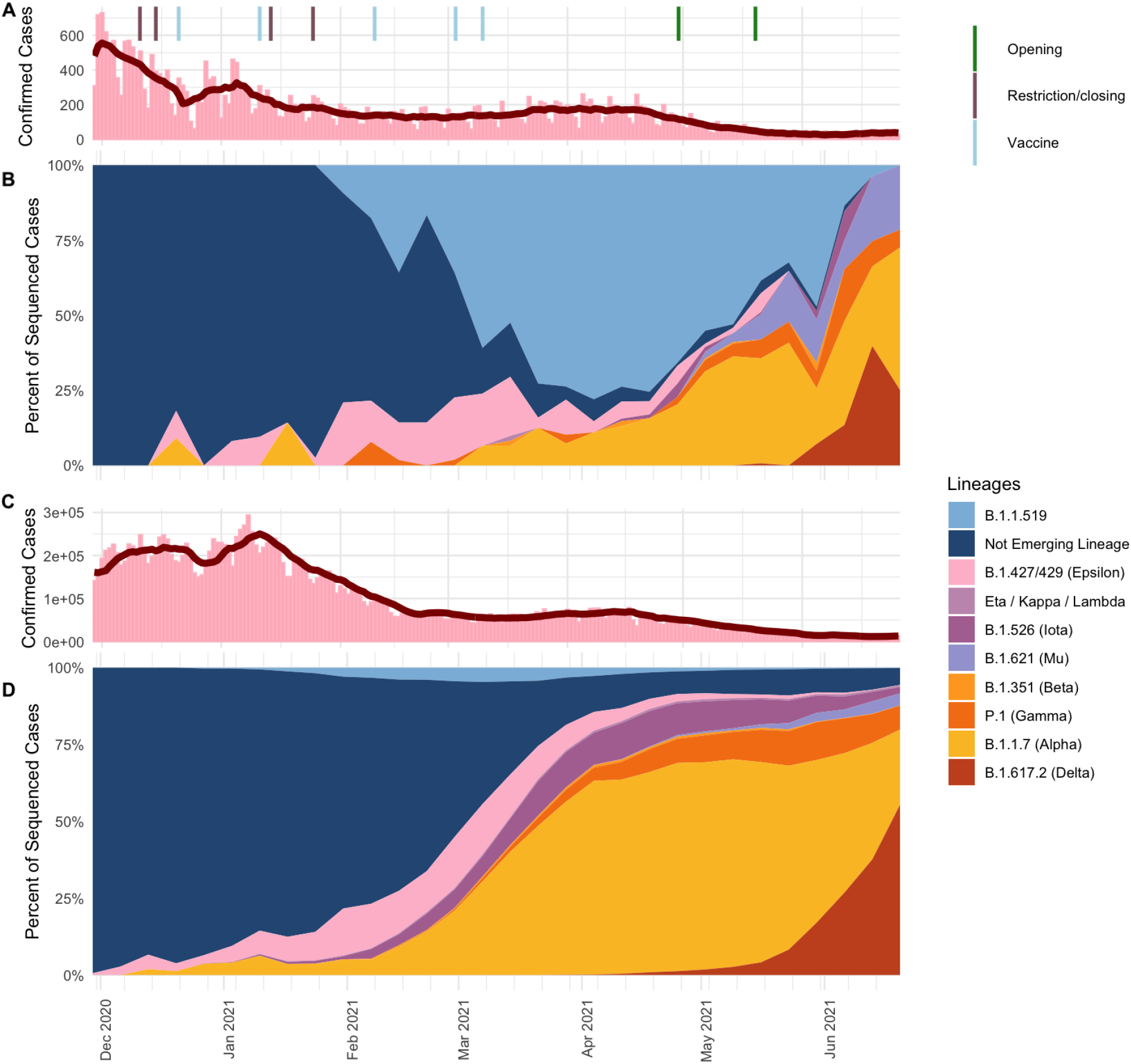
(A and C) Daily case count (bars) and seven day rolling average (red line) and (B and D) percent of sequences (estimated prevalence) by PANGO lineages detected by week from 2020-11-29 to 2021-06-26 for Alaska (A and B) and the Lower 48 (C and D).

Unlike the Lower 48 where Alpha comprised approximately 5% of sequenced cases by the end of January, in Alaska, Alpha was not consistently detected in sequenced cases until March of 2021 (Figure 1B). By the first week of March in Alaska, B.1.1.519 had already reached an estimated 60.8% of sequenced cases compared to 4.6% in the rest of the US, which was the peak prevalence for the Lower 48 (Figure 1B). During B.1.1.519’s peak in the Lower 48, Alpha made up 30.5% of sequenced cases. In Alaska, B.1.1.519 reached a peak prevalence of 77.9% by the week of 4 April 2021, 5 weeks after the peak in the Lower 48.

In terms of COVID-19 cases reported in Alaska, there was a sharp decline the first several weeks of December 2020 followed by an increase in cases through the first week of January 2021 and then another decline (Figure 1A). During this time, cases in the United States increased until the second week of January 2021 followed by a decline through the third week of February 2021 (Figure 1C). In both Alaska and the rest of the United States new COVID-19 cases remained at a relatively stable rate while the cases attributed to VOC, VOI, and VBMs were rising (Figure 1A,C). Given the complexity of factors that affect case rates, it is difficult to attribute the stabilization in cases to biological mechanisms driven by variants or behavioral and social dynamics such as increased vaccination rates, social restrictions, or adherence to respiratory hygiene measures during this time.

When B.1.1.519 first emerged in Alaska, several restrictions were in place as a result of COVID-19 Public Health Disaster Emergency Declaration enacted 11 December 2020 and then renewed 14 January 2021 (Figure 1A). In December, Alaska also received its initial allocations of the Comirnaty vaccine (Pfizer-BioNTech BNT162b2) for frontline workers. By 11 January 2021, Alaskans 65 years of age and older were eligible for both Comirnaty and Spikevax (Moderna mRNA-1273). Vaccine availability was expanded to the entire public by 10 March 2021 and included primarily Spikevax and Comirnaty, at which time B.1.1.519 comprised 60.8% of the sequenced cases. Alaska was ahead of the Lower 48 in terms of vaccine availability and percent of the population vaccinated in early 2021, which confounds the role host immunity may have had in variant emergence and infection dynamics.

### Difference in the emergence of B.1.1.519 within Alaska’s two most populated regions

Given Alaska’s vast geographic expanse and limited travel options, populations of people tend to interact more frequently within economic regions of the state. In Alaska there are six economic regions defined by the Department of Labor and Workforce Development: the Anchorage-Mat Su, Interior, Gulf Coast, Southeast, Southwest, and Northern regions in order from highest to lowest population. Based on available but limited data, we observed distinct trends in SARS-CoV-2 across economic regions of Alaska (Figure S1). However, in our analysis, we excluded all but the two most populous economic regions, the Anchorage-Mat Su and Interior, because of the low proportion of sequenced cases in the other economic regions of the state. We can draw more robust conclusions about estimates of prevalence because these two regions sequenced greater than 5% of the newly reported cases February through June of 2021 (Figure S2).

Between the Anchorage-Mat Su and Interior economic regions there were distinct dynamics in the prevalence of variants over time (Figure 2). For example, Alpha established itself earlier in the Anchorage-Mat Su than the Interior, 360 miles apart. In the Interior, Epsilon comprised a high percentage of sequenced cases from January through March 2021, and the emergence and persistence of B.1.1.519 differed between these two regions of the state.

**Figure 2.**
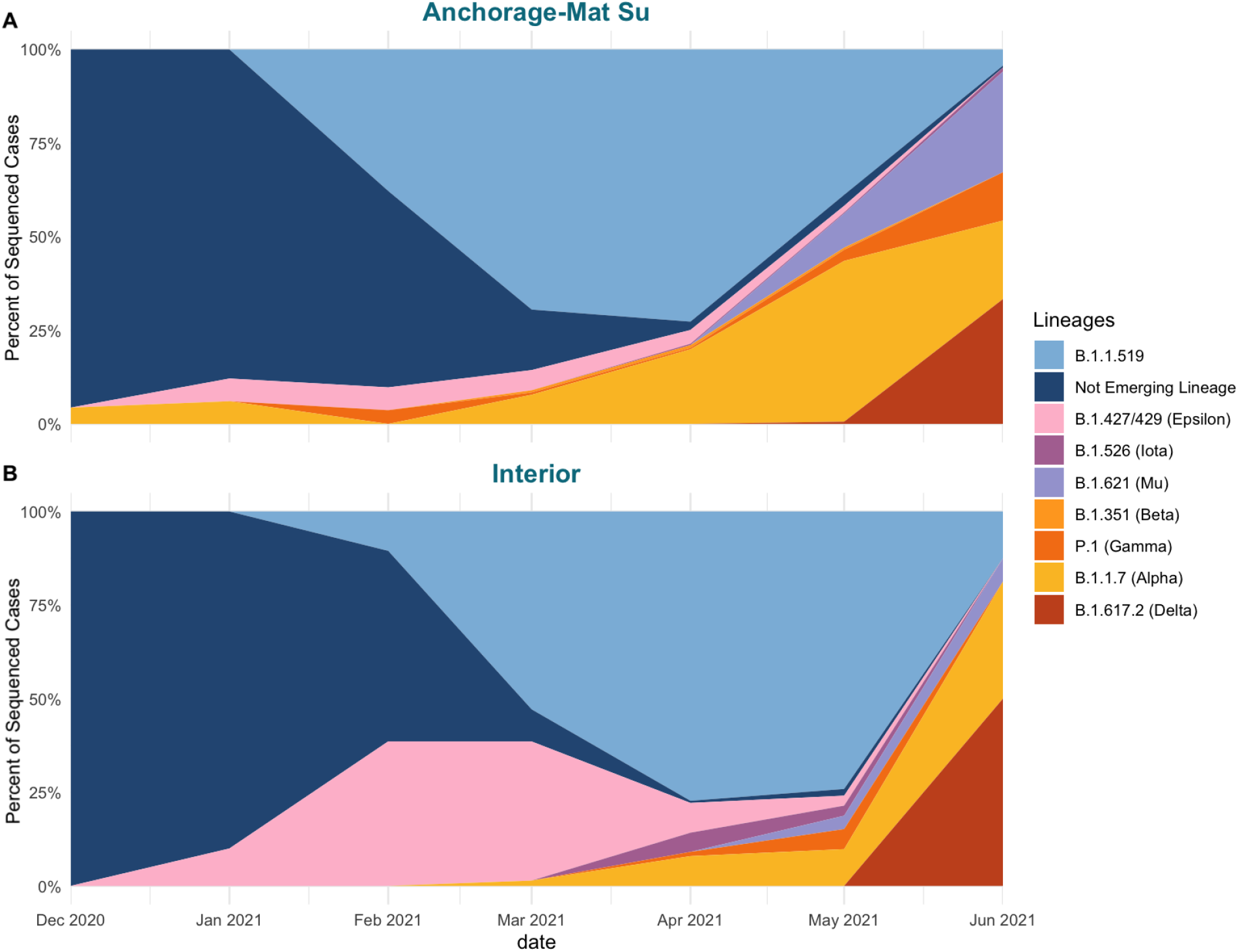
Percent of sequenced cases (estimation of prevalence) by PANGO lineage by month for the (A) Anchorage-Mat Su and (B) Interior regions of Alaska from December 2020 through June 2021.

The first case of B.1.1.519 in Alaska was detected from a sample collected 4 February 2021 in the Southwest region of the state. In both the Anchorage-Mat Su and Interior regions, B.1.1.519 was detected in specimens with collection dates one day apart from one another, 7 and 8 February 2021, respectively. By the end of February, B.1.1.519 comprised 37.8% of sequenced cases in the Anchorage-Mat Su region and 10.5% of cases in the Interior (Figure 2). By February VBMs detected in the Anchorage-Mat Su included Alpha, Epsilon, and Gamma. On the other hand, in the Interior, Epsilon was the only VBM detected and comprised 38.5% of sequenced cases in February (Figure 2). In the Interior, it appears that Epsilon, which was first detected 6 January 2021, was able to establish itself before B.1.1.519. However, one month after B.1.1.519 was first detected in the Interior, B.1.1.519 overtook Epsilon. By April, the majority of sequenced cases were assigned to B.1.1.519 in the Interior (77.2%) and the Anchorage-Mat Su (72.6%). After peaking in April, prevalence of B.1.1.519 in May only marginally decreased for the Interior to 74.1% but sharply decreased for the Anchorage-Mat Su region to 38.8% (Figure 2).

The sharp decline of B.1.1.519 in the Anchorage-Mat Su region may have occurred earlier than in the Interior due to the emergence of other variants that had established themselves in the Anchorage-Mat Su region well before the Interior. For example, in the Anchorage-Mat Su region, Alpha was detected on 20 December 2020 — 97 days before it was first detected in the Interior. This gave Alpha more time to establish itself within the population and consequently reached 42.9% of sequenced cases by May in the Anchorage-Mat Su region versus 9.8% in the Interior. Delta also emerged in the Anchorage-Mat Su region weeks before it was first detected in the Interior, highlighting why B.1.1.519 may have persisted in Interior populations with the last case of B.1.1.519 detected two weeks after last detection in the Anchorage-Mat Su.

### High prevalence of B.1.1.519 in Alaska driven by founder effect and mutational advantage

Mutations in the SARS-CoV-2 genomes from Alaskan cases occurred in an almost clockwise fashion (Figure 3B). Although most of these mutations are expected to have neutral effects, some changes can confer selective advantages that enhance fitness by increasing immune evasion, transmissibility, and/or infectivity (Korber et al., 2020). Mutations that cause amino acid changes to the spike (S) glycoprotein may confer selective advantages given the role of the S protein in COVID-19 pathogenesis and tropism. The mature S protein is cleaved into a fusion domain (S2), and a S1 domain containing an N-terminal head and the receptor binding domain (RBD) that binds to host cell human angiotensin converting enzyme-2 (hACE2) receptor. The S1 domain and RBD particularly are key targets for binding of neutralizing antibodies induced by infection or vaccines, principally IgG (Walls et al., 2020). Given the role of the S protein in receptor binding and subsequent membrane fusion and viral entry into host cells, mutations affecting S1, especially the RBD, can impact hACE2 affinity, viral entry, infectivity, and immune evasion (Berger &Schaffitzel, 2020; Hadi-Alijanvand &Rouhani, 2020).

**Figure 3.**
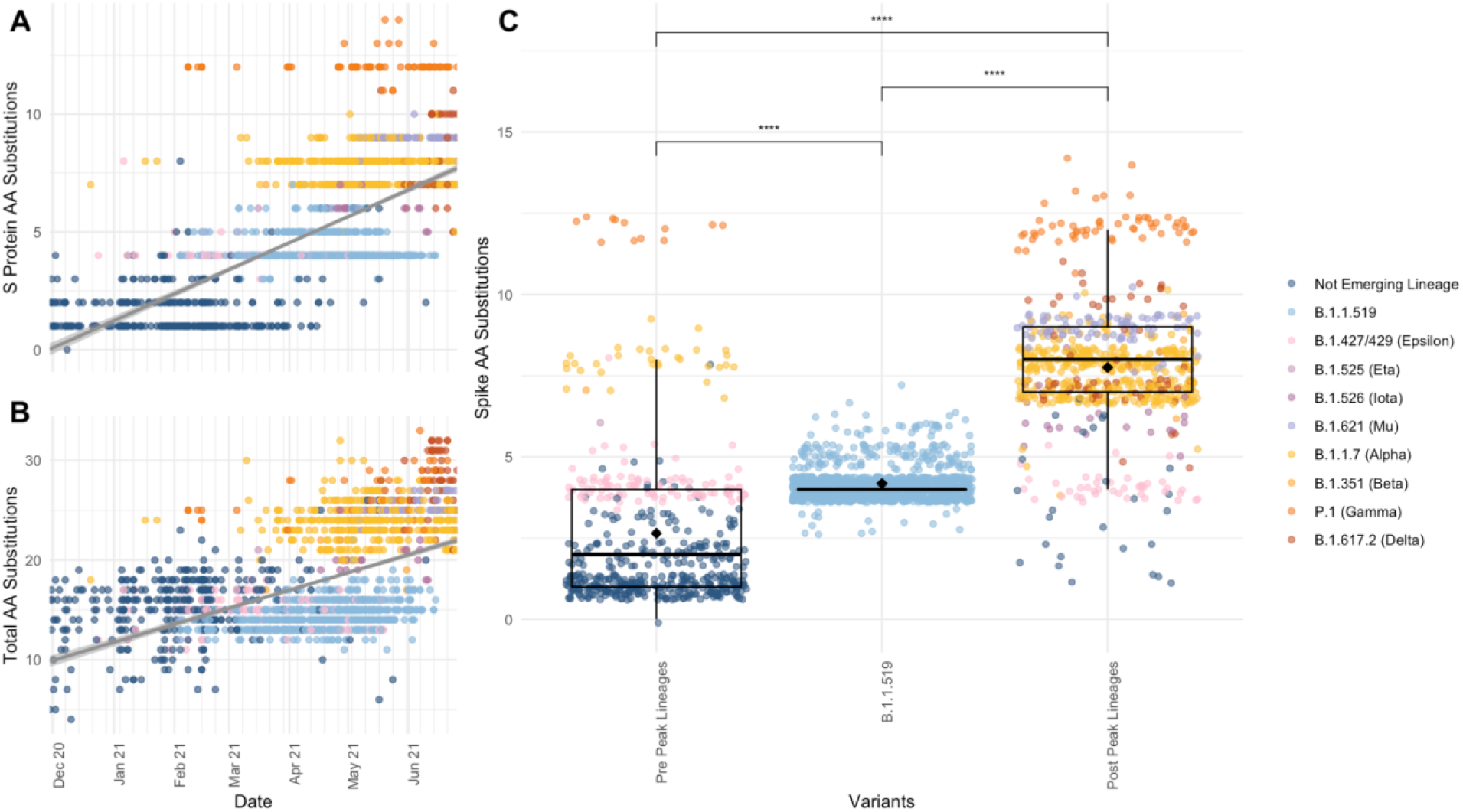
Scatterplots depicting the A) S protein amino acid (AA) substitutions and the (B) total AA substitutions over time. Data includes each genome from an Alaskan confirmed case, colored by variant. The gray line shows a generalized linear model regression of data.(C) Boxplot of number of S protein AA substitutions per genome from sequences pre-peak prevalence of B.1.1.519 on 2021-04-04, B.1.1.519, and post peak prevalence genomes. Diamonds depict the mean number of AA substitutions. Points are individual genomes colored by lineage. Wilcoxon test between group significance *p* < 0.0001 ****, *p* < 0.05 *, ns >0.1.

B.1.1.519 has 11 shared amino acid (AA) mutations relative to the original SARS-CoV-2 (Wuhan-1) genome, with 4 in S: T478K and D614G in the RBD, P681H in the S1/S2 cleavage site, and T732A in the S2 domain (Rodriguez-Maldonado 2021). T478K also arose in B.1.617.2 (Delta) (Jhun, 2021) and may contribute to antibody evasion (Di Giacomo et al., 2021). P681H also occurs in B.1.1.7 (Alpha) and has a weak ability to increase proteolytic cleavage of S1/S2 *in vitro* (Lubinskiet et al., 2021). The functional significance of B.1.1.519 harboring these mutations in S is unclear and in a study from the Netherlands there was no evidence of increased viral load conferred by B.1.1.519 (Gorgels et al., 2022).

Interestingly, based on the regression of total AA changes over time, which exhibited an increase at a steady rate, B.1.1.519 genomes had fewer total AA changes across the genome than predicted (Figure 3A) but aligned with the number of spike protein AA changes (Figure 3B). B.1.1.519 had significantly more AA substitutions in the spike protein than lineages detected in Alaska prior to the week of B.1.1.519’s peak prevalence, 4 April 2021, suggesting a potential competitive advantage over previously circulating lineages (B.1.1.519 = 4.18 ± 0.01, Pre-peak lineages = 2.65 ± 0.10 S AA substitutions, Wilcoxon W = 42302, p-value < 2e-16; Figure 3C). Genomes sequenced after B.1.1.519’s peak prevalence had significantly more AA changes in the spike protein than B.1.1.519, dominated by more divergent Alpha and Delta VOC that emerged after B.1.1.519’s peak (Post-peak lineages = 7.75 ± 0.07 S AA substitutions, Wilcoxon W = 1e+05, p-value < 2e-16; Figure 3C). These VOC likely held a selective advantage that allowed them to outcompete B.1.1.519 in Alaska, a phenomenon of variant replacement observed across Alpha and Delta waves in the USA (Lambrou et al., 2022).

B.1.1.519 established itself within the circulating population of SARS-CoV-2 viruses in Alaska weeks before other variants, such as Alpha, were consistently detected (Figure 1B). The difference in prevalence of B.1.1.519 during Alpha’s emergence in Alaska versus the Lower 48, paired with the finding of more S protein amino acid changes in this variant than previous lineages in Alaska, suggests that in Alaska B.1.1.519 likely emerged and became dominant due to a founder effect. In this context, the founder effect results in reduction of genetic diversity from a founding lineage of SARS-CoV-2 outcompeting the previously circulating lineages because of a selective advantage that the founding population already possessed. The same trend was likely not possible in the other states because Alpha, which proved to outcompete B.1.1.519, had already established itself within the population, outcompeting B.1.1.519 when it emerged. However, other locations displayed similar patterns to Alaska in terms of B.1.1.519’s emergence and spread, most notably Mexico. B.1.1.519 was first detected in Mexico in November 2020, about three months prior to Alaska’s first detection, and was found to rapidly outcompete existing variants to become the dominant variant in the country, comprising 51.5% of sequenced genomes by January 2021 (Rodríguez-Maldonado et al., 2020). By February, 90.9% of Mexico City’s sequenced cases were B.1.1.519. In a study examining the clinical impacts of B.1.1.519 in Mexico City, Cedro-Tanda (2020) found that B.1.1.519-infected patients had a significant increase in adjusted odds ratio for developing severe symptoms of COVID-19 including dyspnea, chest pain and cyanosis, and hospitalizations compared to non-B.1.1.519 infected patients. Unfortunately, we lacked the availability of the necessary data for Alaska to draw conclusions about the clinical impacts of B.1.1.519 in Alaska as shown in the Mexico-City study. Another study, from the Netherlands, describes a cluster of cases associated with B.1.1.519 demonstrating a high within facility attack rate (Gorgels et al., 2022). Interestingly, that same study did not find differences in B.1.1.519 viral loads compared with other variants, and sample size limited conclusions about disease severity.

## Conclusions

Using genomic data, we identified the unique dynamics of B.1.1.519 spread in Alaska compared to the contiguous United States. The distinct pattern of emergence and spread in Alaska emphasizes both how founder events impact regional differences in circulating SARS-CoV-2 lineages and the significance of having robust sequencing efforts in place to detect this variation. There are several resources that compile SARS-CoV-2 sequence data to help examine patterns of emergence and spread. One such resource, outbreak.info, uses genomic data from the sequence repository GISAID to estimate prevalence, emergence, and spread of SARS-CoV-2 lineages globally (Mullen et al., 2020). When considering data from this source, the highest worldwide prevalence of B.1.1.519 was in early March 2021, a month before Alaska’s peak prevalence. At a country level, B.1.1.519 was found to comprise the highest percentage of sequenced genomes in Mexico (15% of sequenced cases compared to 1% of cases sequenced in the United States) over the course of the pandemic. On 14 February 2022, when this analysis was conducted, B.1.1.519 comprised 10.4% of all of Alaska’s SARS-CoV-2 sequences to date. While this is high compared to other geographical locations, spatial and temporal heterogeneity in sequencing efforts is known to bias genomic surveillance and complicate comparisons between locations.

In Alaska, disparate sequencing coverage affected our ability to generate robust analyses in rural regions of the state where sampling and subsequent sequencing efforts were lower than more populated regions. Despite this disparate coverage, genomic surveillance continues to be an essential tool for informing public health decision-making throughout the state. Genomic surveillance paired with epidemiological findings will be key to detecting new threats as they emerge and is a critical tool in shaping policy to address the clinical impacts of local variation in circulating virus lineages. At a patient level, variant surveillance can inform which monoclonal antibodies will be most effective for early treatment to directly reduce individual mortality and morbidity. At a community level, variant proportion data paired with epidemiological findings can inform the broader population of their risks of reinfection, evaluate vaccine efficacy, and assess the need for appropriate treatment stockpiles and quantities. This information will be of great importance for public health policy decisions as SARS-CoV-2 becomes endemic in Alaska and around the globe.

## Data Availability

All data used in this study are available online at GISAID (https://www.gisaid.org/) and NCBI GenBank (https://www.ncbi.nlm.nih.gov/sars-cov-2/). Metadata on the number of cases for Alaska is available on the Alaska COVID-19 Summary Dashboard (https://covid19.alaska.gov/)

## Data Availability Statement

All data used in this study are available online at GISAID (www.gisaid.org/) and NCBI GenBank (www.ncbi.nlm.nih.gov/sars-cov-2/).

## Acknowledgments

We gratefully acknowledge the researchers responsible for obtaining the specimens and the laboratories where genetic sequence data were generated and shared via the GISAID Initiative and NCBI GenBank, on which this research is based.

## Supplemental Figures

**Figure S1.**
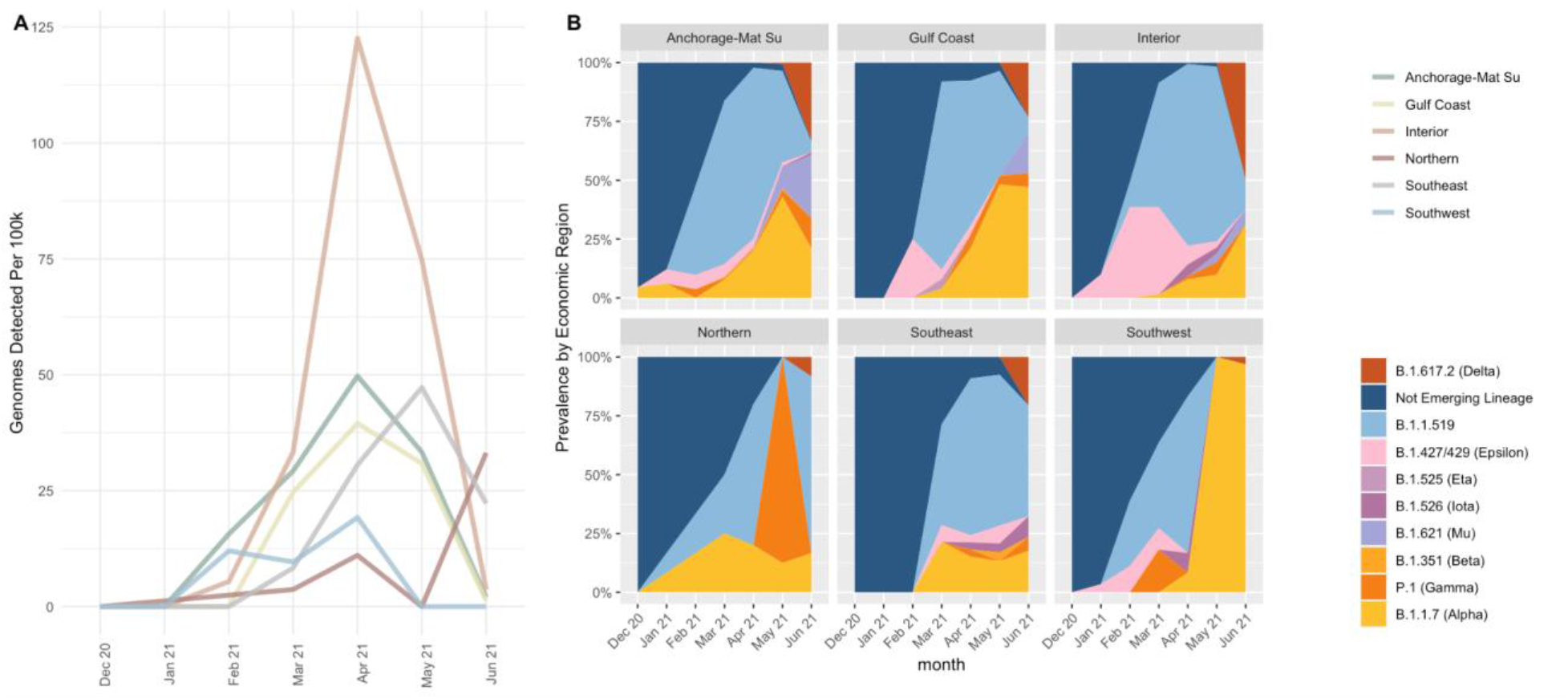
(A) Number of detected B.1.1.519 genomes per 100k people by Alaska economic region over time. Lines are colored by Alaska economic region. (B) Estimated prevalence of emerging lineages by Alaska economic region from December 2020 through June 2021.

**Figure S2.**
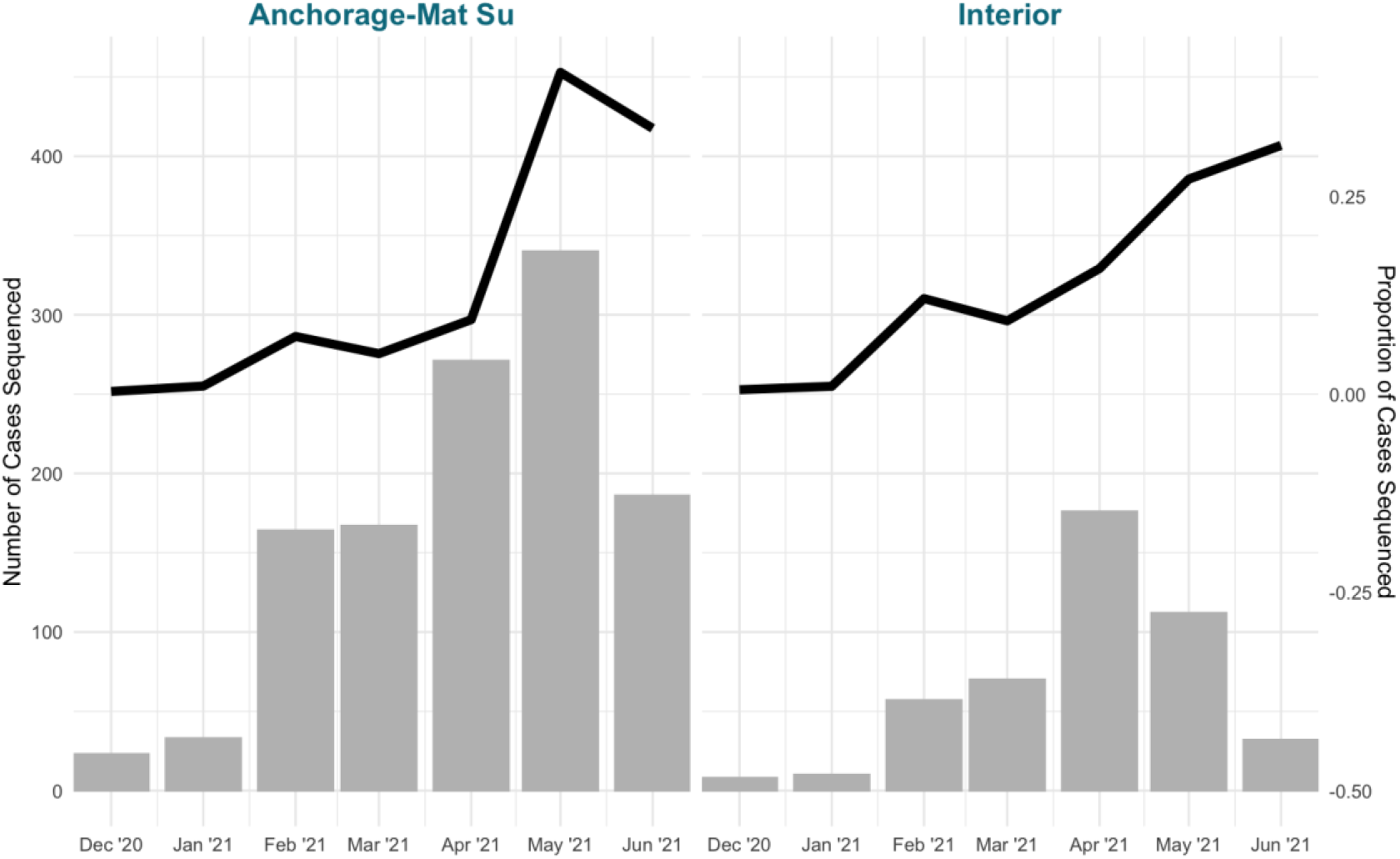
Number of cases sequenced (bars) and the proportion of total cases sequenced in the Anchorage-Mat Su and Interior economic regions of Alaska.

